# Fairness-aware, explainable clinical decision support for opioid use disorder risk stratification: development and internal validation of a dual-layer AI system

**DOI:** 10.64898/2026.06.23.26356401

**Authors:** Mehmet Kazgan, Navid Mohammadvand, Burak Cetin

## Abstract

Opioid use disorder (OUD) remains a leading cause of preventable death in the United States, yet the tools used to assess OUD risk rely on episodic self-report, produce binary output, and exhibit documented performance disparities across demographic groups that can widen existing inequities in care. Machine-learning models for OUD risk are seldom evaluated for demographic fairness or designed for the transparency clinicians need to trust and act on them. We present a fairness-aware, explainable clinical decision support system for four-tier OUD risk stratification, developed and internally validated on a large electronic health record–derived cohort accessed via Mayo Clinic Platform_Discover. The system pairs an XGBoost classifier with a transparent Clinical Rules Engine that attributes risk across six clinical domains, providing clinician-interpretable explanations alongside each prediction. To address demographic disparity directly, we applied an iterative bias-mitigation strategy combining age-balanced resampling, removal of race as a model input, and cost-sensitive reweighting, and measured its effect using group-fairness metrics (demographic parity, equal opportunity, equalized odds, and calibration within groups). On a held-out internal test set, mitigation reduced the White–Black gap in high-risk detection from 30.3 to 7.4 percentage points (a 76% relative reduction) and the age-based accuracy gap from 6.6 to 2.7 percentage points (59% reduction), raising high-risk detection for Black patients from 58.3% to 75.0%, at a cost of fewer than two percentage points of overall accuracy; gender differences remained below three points. The system was independently qualified through Mayo Clinic Platform_Solutions Studio. This work offers an implementable, transparent blueprint for operationalizing fairness and explainability in clinical AI for high-risk prescribing, with external and prospective validation as the clear next steps.

## Background and significance

The opioid crisis in the United States represents one of the most devastating public health challenges of the past three decades. From 1999 through 2023, opioid-related overdose deaths exceeded 600,000 [1,2], with synthetic opioids accounting for an increasing share of fatalities. The economic burden exceeds $1 trillion annually [3] when accounting for healthcare costs, lost productivity, criminal justice involvement, and social services. Despite substantial public health investments, early identification of patients at risk for developing opioid use disorder (OUD) before addiction takes hold remains a critical unmet need.

Current clinical approaches to OUD risk assessment rely on a fragmented combination of screening questionnaires, provider judgment, and manual review of prescription drug monitoring program (PDMP) data. The most widely used instruments—the SOAPP-R and the ORT—achieve 68–77% and 65–75% accuracy [4,5], respectively, in published validation studies. These tools share fundamental limitations: they depend on episodic self-reported data, provide binary rather than graded risk assessment, require separate clinical workflows, and identify risk only after problematic patterns have already emerged. Clinical judgment alone achieves approximately 50–60% accuracy [6], with substantial inter-provider variability.

Machine learning approaches to OUD risk prediction have shown promise in research settings, with several studies demonstrating that electronic health record (EHR) data can support predictive modeling for substance use disorders [7–9]. However, significant barriers to clinical adoption remain. Published models frequently lack explainability, raising concerns about clinician trust and regulatory acceptability. Fairness evaluation is often absent or retrospective, with documented demographic performance disparities in healthcare AI systems [10]. Perhaps most critically, few models have progressed beyond retrospective validation [21] to qualification on clinical platforms that support real-world deployment.

### Objective

To address these gaps, we developed cliexaAI, a dual-layer clinical decision support system that combines machine learning prediction with rule-based clinical reasoning to provide four-tier OUD risk stratification. The system was developed using de-identified data accessed via Mayo Clinic Platform_Discover and qualified through Mayo Clinic Platform_Solutions Studio. This paper describes the system architecture, iterative bias mitigation methodology, performance evaluation, and qualification process, with emphasis on three contributions to the field:

- A dual-layer architecture pairing an XGBoost classifier with a Clinical Rules Engine (CRE) that provides transparent, clinician-interpretable risk factor attribution alongside machine learning prediction.
- A documented and measured bias mitigation strategy that achieved 76% reduction in racial performance disparity and 59% reduction in age-based disparity through iterative dataset rebalancing, feature selection, and fairness-aware training.
- Qualification on Mayo Clinic Platform as a production-ready clinical decision support tool, establishing a pathway for responsible deployment of AI-driven OUD prevention.

This work is positioned within the domain of clinical decision support (CDS), with a focus on integrating explainable AI into electronic health record workflows to support real-time risk assessment and clinician decision-making.

## Materials and methods

### Study design and ethical approval

This study involves analysis of de-identified data via Mayo Clinic Platform_Discover. In accordance with the Code of Federal Regulations, 45 CFR 46.102, the noted activity does not require Institutional Review Board (IRB) review [11].

Data shown and reported in this manuscript has been extracted from this environment using an established protocol for data extraction, aimed at preserving patient privacy. The data has been de-identified pursuant to an expert determination in accordance with the HIPAA Privacy Rule. Any data beyond what is reported in the manuscript, including but not limited to the raw data, cannot be shared or released due to the parameters of the expert determination to maintain data de-identification.

The study followed a retrospective model development design using structured clinical data, followed by prospective qualification through Mayo Clinic Platform_Solutions Studio. Three model versions were developed iteratively between September and December 2025, with each version incorporating findings from bias analysis and fairness evaluation. The system is designed for integration into EHR-based clinical workflows; however, real-time clinical deployment and outcome evaluation are ongoing.

### Data source and patient population

The predictive modeling framework was developed using structured, de-identified clinical data accessed via Mayo Clinic Platform_Discover. Data elements included ICD-10 coded diagnoses, prescription records, laboratory values, patient encounters, demographic information, medication history, behavioral health records, and patient-reported outcomes (PROs) including PHQ-9, GAD-7, and SOAPP assessments where available. Data extraction was performed using SQL queries with mappings across tables via anonymized patient identifiers and standardized lookup joins for diagnosis codes and clinical service classifications.

#### Baseline population

Approximately 204,000 patients were identified through opioid prescription records with valid inclusion and exclusion filters. Inclusion criteria required: age 18–80 years, at least one opioid prescription (including oxycodone, hydrocodone, morphine, fentanyl, hydromorphone, oxymorphone, tapentadol, codeine, or tramadol), valid ICD-10 diagnoses, and a minimum 12-month observation period with proper temporal sequencing between prescriptions, diagnoses, and outcomes.

Exclusion criteria encompassed: patients with active cancer diagnoses (ICD-10: C00–C96, D37–D48), palliative care (ICD-10: G89.3, Z51.5, Z85), medication-assisted treatment (MAT) without concurrent opioid prescriptions, life-threatening emergencies, post-operative pain management, emergency department crisis care, and records with medication timeline inconsistencies.

#### Cohort assignment

Patients meeting inclusion criteria were assigned to four sub-cohorts with the following prioritization hierarchy: Confirmed OUD > Addiction Treatment Seekers > Overdose Outcome > Baseline Opioid Users. Patients with overlapping inclusion criteria were assigned to the highest-priority cohort. SQL filtering pipelines included iterative cohort count checks, exclusion logic, and cross-validation. Deduplication logic ensured no repeated patient records across model cohorts.

#### Final model development datasets

Two datasets were developed across the study’s iterative model versions:

##### Original dataset (V1, V2)

40,000 patients with complete risk assessments, split into training (32,000; 80%), validation (4,000; 10%), and test (4,000; 10%) sets with stratification by risk tier.

##### Bias-mitigated balanced dataset (V3)

Following identification of demographic performance disparities, a balanced sampling approach reduced the dataset to 15,713 patients: training (12,571; 80%), validation (1,571; 10%), and test (1,571; 10%). The full cohort derivation is shown in Fig 1.

**Fig 1.**
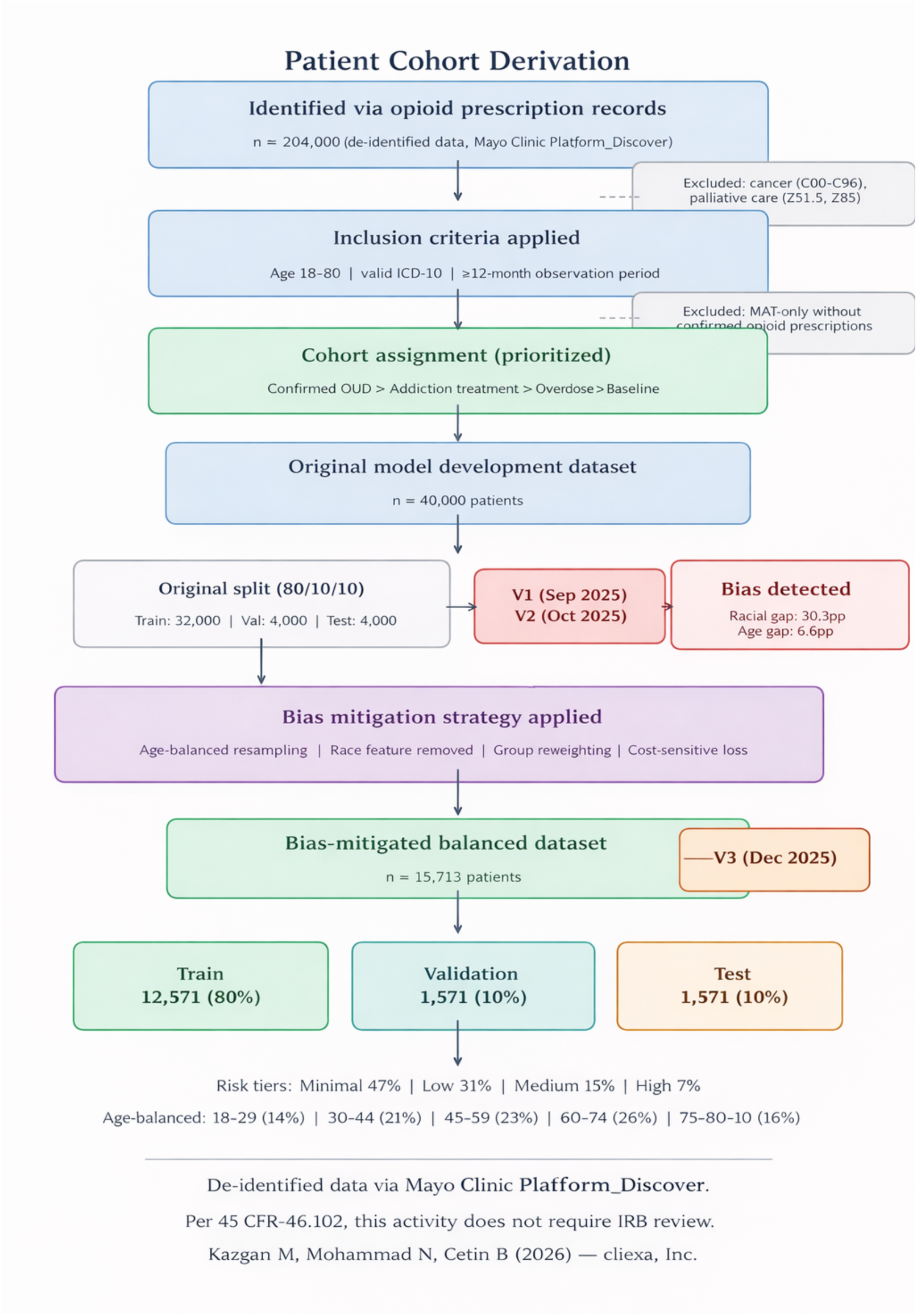
CONSORT-style patient cohort flow diagram. The derivation path from ∼204,000 initial patients identified via opioid prescription records in de-identified data from Mayo Clinic Platform_Discover, through inclusion/exclusion filtering, cohort assignment, original 40,000-patient dataset (V1/V2), bias detection, and bias mitigation strategy (age-balanced resampling, race feature removal) to the final 15,713-patient balanced dataset (V3) with 80/10/10 train/validation/test split.

The balanced dataset achieved the following age distribution: 18–29 (14.0%), 30–44 (21.0%), 45–59 (23.0%), 60–74 (26.0%), 75–80 (16.0%). Risk tier distribution: Minimal (47.0%), Low (30.6%), Medium (15.4%), High (7.0%). Gender distribution was approximately 54.6% male and 45.3% female (Table 1).

**Table 1.** Balanced dataset demographics and risk tier distribution (n=15,713).

| Age group | Train | Test | % (Train) | Risk tier | % Total |
| --- | --- | --- | --- | --- | --- |
| 18–29 | 1,760 | 220 | 14.0% | Minimal | 47.0% |
| 30–44 | 2,640 | 330 | 21.0% | Low | 30.6% |
| 45–59 | 2,891 | 361 | 23.0% | Medium | 15.4% |
| 60–74 | 3,269 | 408 | 26.0% | High | 7.0% |
| 75–80 | 2,011 | 252 | 16.0% |  |  |
| Total | 12,571 | 1,571 | 100% | Total | 100% |

### Feature engineering and preprocessing

Approximately 42 structured features were engineered across six clinical domains aligned with the Clinical Rules Engine architecture: event history risk, medications, laboratory data, comorbidities, social score, and patient-reported outcomes.

#### Medication history features

Current versus past-only opioid status indicators, days since last opioid prescription, morphine milligram equivalent (MME) calculations, provider and pharmacy diversity scores (COUNT DISTINCT within 90-day windows), and fill/refill patterns.

#### Comorbidity encoding

Conditions including depression, PTSD, anxiety, and COPD encoded as ordinal variables based on clinical severity, with features derived from ICD-10 code groupings.

#### PRO integration

Binary indicators of severity thresholds (e.g., PHQ-9 > 9 indicating positive screen), with planned composite score calculations for anxiety–depression interactions.

#### Behavioral engagement

Frequency of addiction-related encounters, number of unique visit types and behavioral specialists seen, and appointment duration patterns.

#### PDMP indicators

Count of PDMP risk flags used as risk multipliers within the feature set.

Categorical variables were encoded using one-hot encoding for non-ordinal variables (sex, region, diagnosis categories) and ordinal encoding for hierarchical variables (smoking history, PRO severity levels). Risk tier labels were encoded as integers (0=Minimal, 1=Low, 2=Medium, 3=High). Continuous features were left unscaled for the XGBoost pipeline given the algorithm’s native handling of raw values in tree-based splits.

#### Race feature removal

Race was removed from the model feature set as part of the bias mitigation strategy implemented in V3. Race data is retained exclusively in metadata for post-hoc fairness auditing and longitudinal monitoring purposes. This decision was made to eliminate direct demographic bias in predictions while preserving the ability to evaluate model equity across racial groups.

### Missing data handling

Overall missingness varied by data domain: PROs (∼12–15%), laboratory values (∼10%), behavioral history (∼5–8%), medication records (∼2%), and social scores/engagement metrics (up to 18% in some cohorts). Categorical features used an explicit “missing” category to preserve potential informativeness. Continuous features were imputed using median values. XGBoost’s native missing data handling via learned default split directions was leveraged as the primary strategy, with explicit missing indicators provided as supplementary features.

### Model architecture: dual-layer prediction system

#### Layer 1: XGBoost multi-class classifier

The machine learning layer employs an XGBoost multi-class classifier configured with the multi:softprob objective function, which outputs calibrated class probabilities for each of the four risk tiers. The model was implemented using XGBoost v1.6 [12] on a cloud-based instance (6-core CPU, 38 GB RAM, 150 GB disk). The final hyperparameter configuration is provided in S1 Table.

The hyperparameter evolution from V1 to V3 reflects a deliberate regularization strategy: reducing max_depth (7→5), increasing learning_rate (0.01→0.03), and reducing subsample (0.85→0.75) to build a more generalizable model on the smaller, balanced dataset. Training runtime was approximately 30 minutes per full cycle.

#### Layer 2: Clinical Rules Engine (CRE)

The Clinical Rules Engine serves as the foundational logic layer, integrating structured clinical knowledge, weighted heuristics, and dynamic rule evaluation. The CRE evaluates patient risk across six core data buckets: event history risk, medications, laboratory data, comorbidities, social score, and patient-reported outcomes. The overall dual-layer system architecture is summarized in Fig 2.

**Fig 2.**
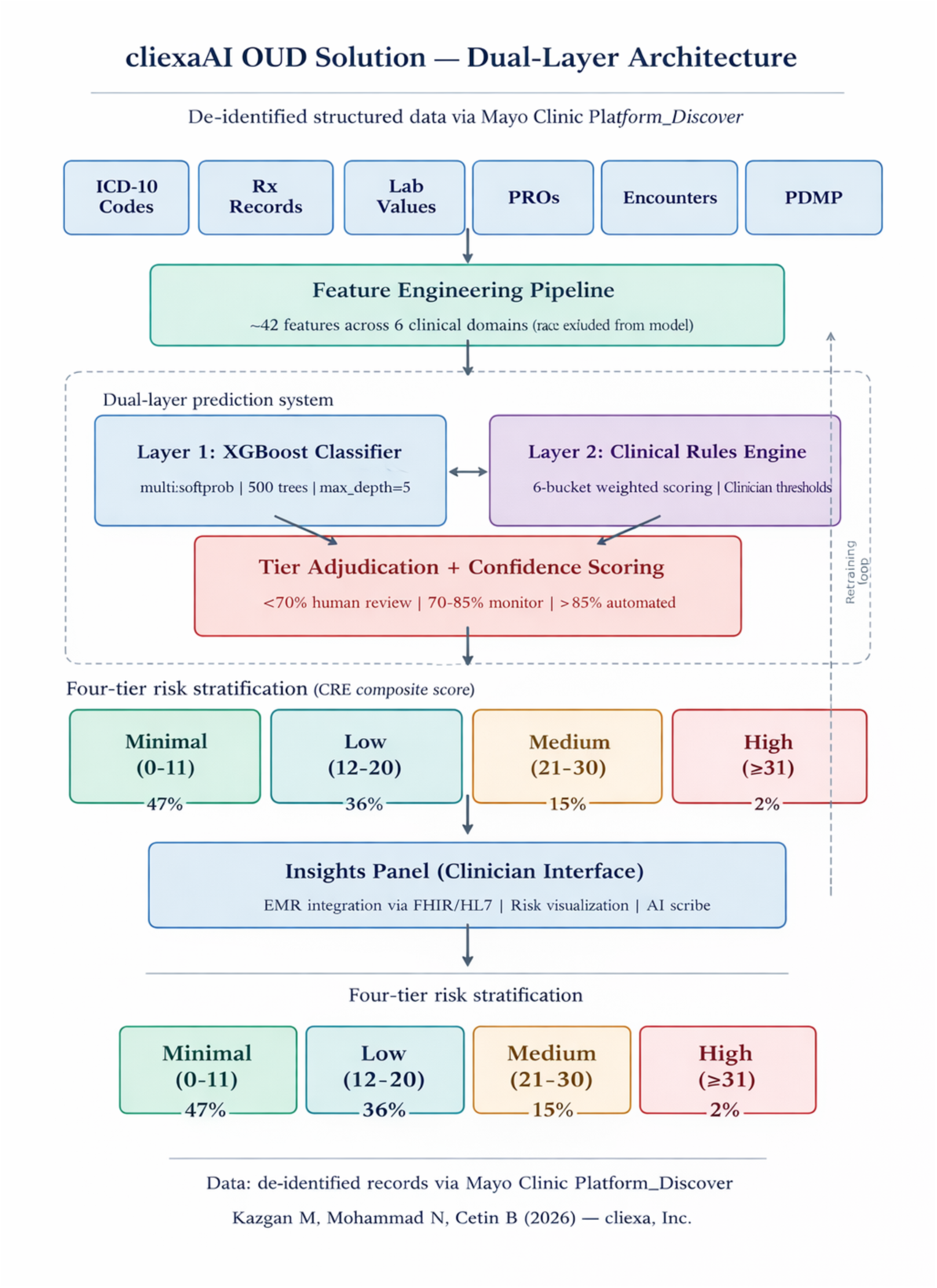
System architecture of the cliexaAI OUD dual-layer clinical decision support system. De-identified structured data from Mayo Clinic Platform_Discover is processed through a feature engineering pipeline (∼42 features across 6 clinical domains, with race excluded from model input). Layer 1 (XGBoost multi-class classifier) and Layer 2 (Clinical Rules Engine with clinician-derived tier boundaries) jointly produce four-tier risk stratification through confidence-based adjudication. Risk output is delivered through a clinician-facing Insights Panel with EMR integration.

Each bucket independently contributes a weighted risk score based on predefined thresholds and severity scales. Component scores are aggregated to form a composite OUD risk profile with the following tier boundaries (clinician-derived, version-controlled): Minimal (0–11 points), Low (12–20 points), Medium (21–30 points), High (≥31 points). Weighted priority ensures that high-significance indicators (e.g., high MME values, positive overdose history) are weighted more heavily than lower-acuity risk factors.

The CRE handles data completeness through a two-pronged system: “No Data” (client lacks access to data element; scored as 0 points) versus “No History” (patient has no relevant history; scored as 0 points with no flag). Required data elements trigger computation halts if missing.

#### Tier adjudication and confidence scoring

The dual-layer system adjudicates between ML prediction and CRE output through a confidence-based triage framework: predictions with confidence below 70% require mandatory human review (active human-in-the-loop); confidence between 70–85% triggers enhanced monitoring (passive human-in-the-loop); confidence above 85% proceeds with automated classification under clinical oversight. A single global threshold is applied across all demographic groups no subgroup-specific cut-offs are used.

### Bias mitigation strategy

The bias mitigation strategy was developed iteratively across three model versions in response to systematic fairness evaluation:

#### V1 (September 2025)

Initial submission with both XGBoost and multi-layer perceptron (MLP) architectures on 40,000 patients. Fairness analysis revealed a 30.3 percentage-point gap in high-risk detection accuracy between White (88.6%) and Black (58.3%) patients, and a 6.6 percentage-point accuracy gap between younger (18–44; 80.2%) and older (60–80; 86.8%) adults.

#### V2 (October 2025)

Added comprehensive subgroup analysis stratified by age, gender, race, medication type, PDMP indicators, and composite risk profiles. The MLP narrowed the racial gap to 23.4 percentage points but did not eliminate it.

#### V3 (December 2025)

Implemented the full bias mitigation strategy and submitted for qualification. The V3 bias mitigation strategy comprised four components:

- Age-balanced resampling: The dataset was reduced from 40,000 to 15,713 patients using constrained sampling to achieve fairer age representation (reducing the 60–80 age group from 50.2% to 42.0% while increasing the 18–29 group from 5.5% to 14.0%).
- Race feature removal: Race was removed as a model input feature to eliminate direct demographic bias. Race is retained in metadata exclusively for post-hoc fairness auditing.
- Group reweighting with cost-sensitive loss: Applied to reduce high-risk false negatives in under-represented groups during training.
- Subgroup calibration: TPR/FPR deltas audited on validation and test sets across demographic groups.

The MLP architecture was deliberately excluded from the final submission despite higher raw accuracy (90.5% vs. 84.2%) because: (a) the MLP achieved only modest racial gap reduction (30.3→23.4 percentage points versus XGBoost’s 30.3→7.4 with balanced sampling), (b) XGBoost provides native feature importance metrics essential for CRE integration, and (c) XGBoost’s tree-based architecture supports the transparency requirements of clinical deployment.

### Evaluation metrics

Model performance was evaluated using overall accuracy, Cohen’s Kappa (κ), Matthews Correlation Coefficient (MCC), macro-averaged and weighted precision, recall, and F1-score, per-tier confusion matrices, and multi-class log loss (mlogloss). Fairness was assessed using Demographic Parity, Equal Opportunity, Equalized Odds, and Calibration within Groups, computed via the Fairlearn toolkit [14]. Subgroup performance was stratified by age group, gender, and race. These metrics operationalize the Fairness principle of the FUTURE-AI international consensus guideline for trustworthy and deployable clinical AI [22], under which an equitable model should maintain comparable performance across patient subgroups. Accordingly, we report subgroup performance parity—the disparity in per-tier detection and accuracy between demographic groups—as a primary outcome, measured before and after mitigation.

### Qualification process

The solution was qualified through Mayo Clinic Platform_Solutions Studio [20], which evaluates solutions’ claims and supporting documentation to provide transparent insight into intended use, proposed value, and clinical and algorithmic performance. The qualification process assessed adherence to responsible AI principles, clinical validity of risk tier definitions, documentation of training methodology and performance metrics, bias evaluation and mitigation strategies, and intended use/out-of-scope use case definitions.

## Results

### Overall model performance

The final bias-mitigated XGBoost model (V3) was evaluated on the held-out test set (n=1,571). Overall performance metrics are summarized in Table 2.

**Table 2.** Overall model performance: original versus bias-mitigated model.

| Metric | Original (Train) | Balanced (Train) | Original (Test) | Balanced (Test) |
| --- | --- | --- | --- | --- |
| Overall Accuracy | 86.7% | 84.1% | 84.2% | 82.3% |
| Cohen's Kappa | 0.792 | 0.756 | 0.757 | 0.729 |
| MCC | 0.785 | 0.757 | 0.730 | 0.730 |
| Macro-F1 | 84.3% | 79.9% | 79.9% | 79.5% |
| Weighted-F1 | 86.3% | 84.0% | 82.2% | 82.2% |

### Per-tier classification performance

As shown in Table 3, the Minimal tier demonstrated the strongest performance (F1 = 90.1%), correctly identifying the majority of patients without significant OUD risk. The High tier achieved precision of 82.7% and recall of 78.2%, supporting targeted intervention use cases. The Medium tier showed the lowest precision (69.5%), reflecting the inherent clinical difficulty of distinguishing moderate-risk patients from adjacent tiers. The model converged after approximately 500 estimators, with multi-class log loss decreasing from 1.4 to approximately 0.45 (training) and 0.55 (validation).

**Table 3.** Per-class test set performance (bias-mitigated model, n=1,571).

| Risk tier | Precision | Recall | F1-score | Support (n) |
| --- | --- | --- | --- | --- |
| Minimal | 88.6% | 91.6% | 90.1% | 738 |
| Low | 78.9% | 73.0% | 75.8% | 481 |
| Medium | 69.5% | 74.4% | 71.9% | 242 |
| High | 82.7% | 78.2% | 80.4% | 110 |

### Confusion matrix analysis

The test set confusion matrix for the bias-mitigated model is provided in S2 Table, and a visual comparison against the original model is shown in Fig 3.

**Fig 3.**
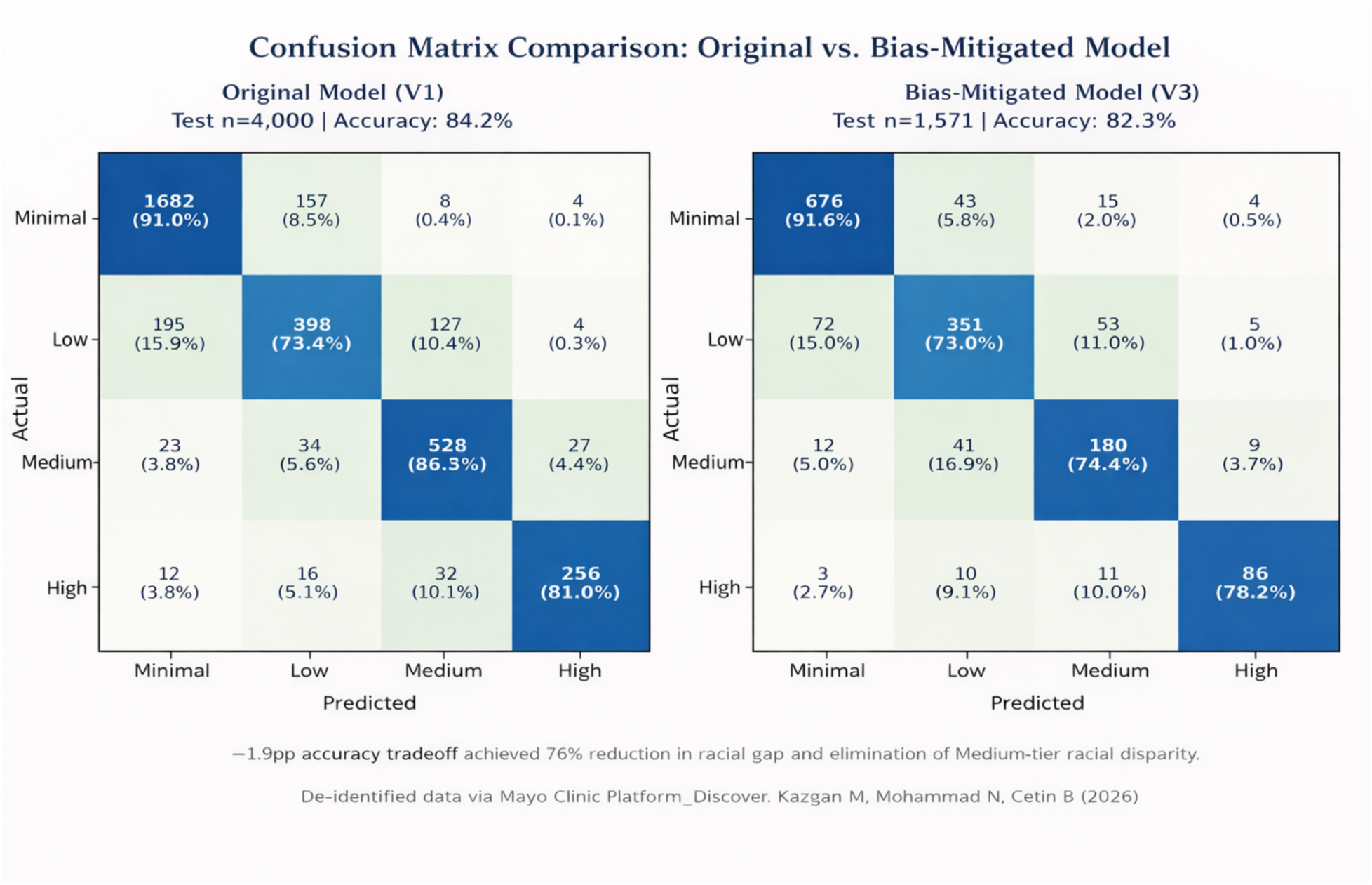
Side-by-side confusion matrix comparison. Left: original model (V1, test n=4,000, accuracy 84.2%). Right: bias-mitigated model (V3, test n=1,571, accuracy 82.3%). The 1.9pp accuracy tradeoff achieved 76% reduction in the racial performance gap and effective elimination of the Medium-tier racial disparity.

Misclassification patterns demonstrate clinically appropriate behavior: errors predominantly occur between adjacent risk tiers rather than distant ones. Only 0.5% of Minimal-risk patients were misclassified as High, and only 2.7% of High-risk patients were misclassified as Minimal. The most common misclassification was Low-risk patients classified as Minimal (15.0%), representing a conservative error pattern.

### Fairness analysis: bias mitigation results

Table 4 summarizes the impact of the bias mitigation strategy across demographic dimensions. Across all three subgroup dimensions, mitigation moved the model toward the equal-performance criterion of the FUTURE-AI Fairness principle [22]. Subgroup performance parity for high-risk detection improved substantially: the White–Black detection gap narrowed from 30.3 to 7.4 percentage points (a 76% relative reduction), and the age-based accuracy gap narrowed from 6.6 to 2.7 percentage points (59% relative reduction), while gender-based F1 differences remained below 3 percentage points throughout. In equalized-odds terms, the largest residual disparity—between true-positive rates for White and Black patients in the high-risk tier—was reduced by roughly three-quarters at a cost of 1.9 percentage points of overall test accuracy.

**Table 4.** Bias mitigation results: before (V1) versus after (V3) mitigation.

| Dimension | Before | After | Change (pp) | % Reduction |
| --- | --- | --- | --- | --- |
| Racial gap: high-risk (White vs. Black) | 30.3pp | 7.4pp | −22.9 | 76% |
| Age gap (18–44 vs. 60–80) | 6.6pp | 2.7pp | −3.9 | 59% |
| Gender gap (F1 across tiers) | <3% | <3% | Maintained | N/A |
| Black high-risk accuracy | 58.3% | 75.0% | +16.7 | — |
| Overall accuracy cost (test) | 84.2% | 82.3% | −1.9 | — |

Per-tier racial gap analysis showed the most dramatic improvement in the Medium tier, where the gap was effectively eliminated (13.0pp → −1.0pp, with Black patients marginally outperforming White patients at 75.9% vs. 74.9%). The High tier gap was reduced from 30.3pp to 7.4pp. Age-based accuracy variance (σ) decreased from 2.63% to 1.13% across age groups, representing a 57% reduction in performance variability. These before-and-after fairness outcomes are visualized in Fig 4.

**Fig 4.**
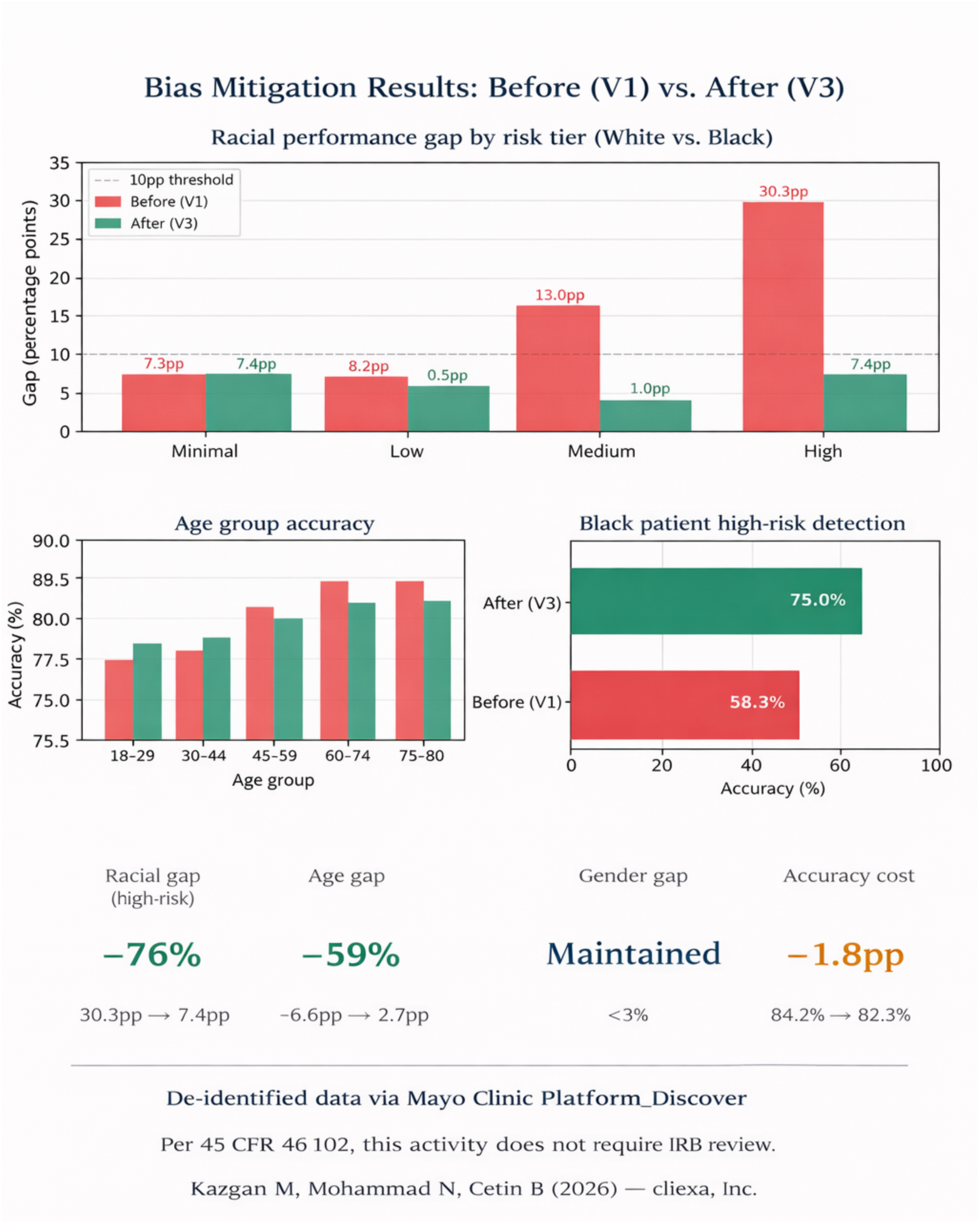
Bias reduction dashboard showing before-and-after fairness metrics. Top: summary metric cards for racial gap reduction (76%), age gap reduction (59%), gender equity maintenance (<3%), and accuracy cost (1.8pp). Charts: (a) racial performance gap by risk tier comparing original and bias-mitigated models, (b) age group accuracy before and after rebalancing, (c) Black patient high-risk detection accuracy improvement from 58.3% to 75.0%.

### Feature importance

The top predictive features based on gain importance in the bias-mitigated model were: drug-related disorder diagnosis (18.2%), filled prescription count (14.1%), depression severity (11.8%), appointment duration (9.7%), MME (8.9%), urinalysis results (7.2%), and age (6.1%) (Fig 5). These features align with established clinical risk factors for OUD and support the interpretability of model predictions. SHAP-based per-patient explainability analysis is planned for the next iteration to provide individualized risk factor attribution.

**Fig 5.**
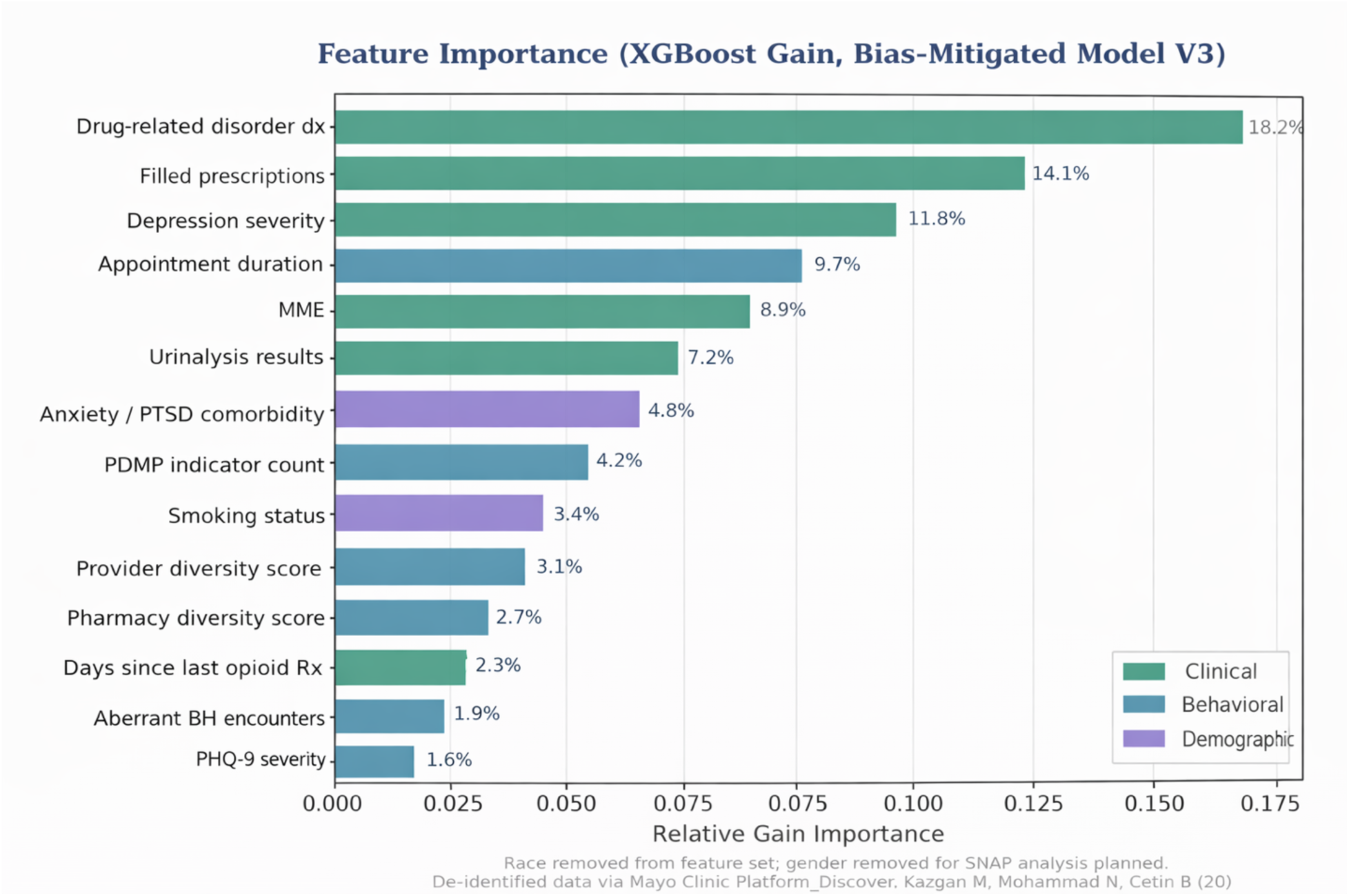
Feature importance based on XGBoost gain metric (bias-mitigated model, V3). Top 15 features ranked by relative gain importance, color-coded by domain: clinical (green), behavioral (blue), and demographic (purple). Drug-related disorder diagnosis (18.2%) and filled prescription count (14.1%) are the strongest predictors. Race was removed from the feature set as part of bias mitigation.

### Comparative context

While direct head-to-head comparison is not possible due to differences in study populations, outcome definitions, and evaluation methodologies, the cliexaAI system’s 82.3% test accuracy provides useful context against published performance ranges of existing screening tools (Fig 6). The SOAPP-R achieves 68–77% sensitivity in prospective validation studies using 24-item self-report questionnaires [4,15]. The ORT achieves 65–75% accuracy with a 5-item self-report instrument [5,16]. Clinical judgment alone is estimated at 50–60% accuracy with substantial inter-provider variability.

**Fig 6.**
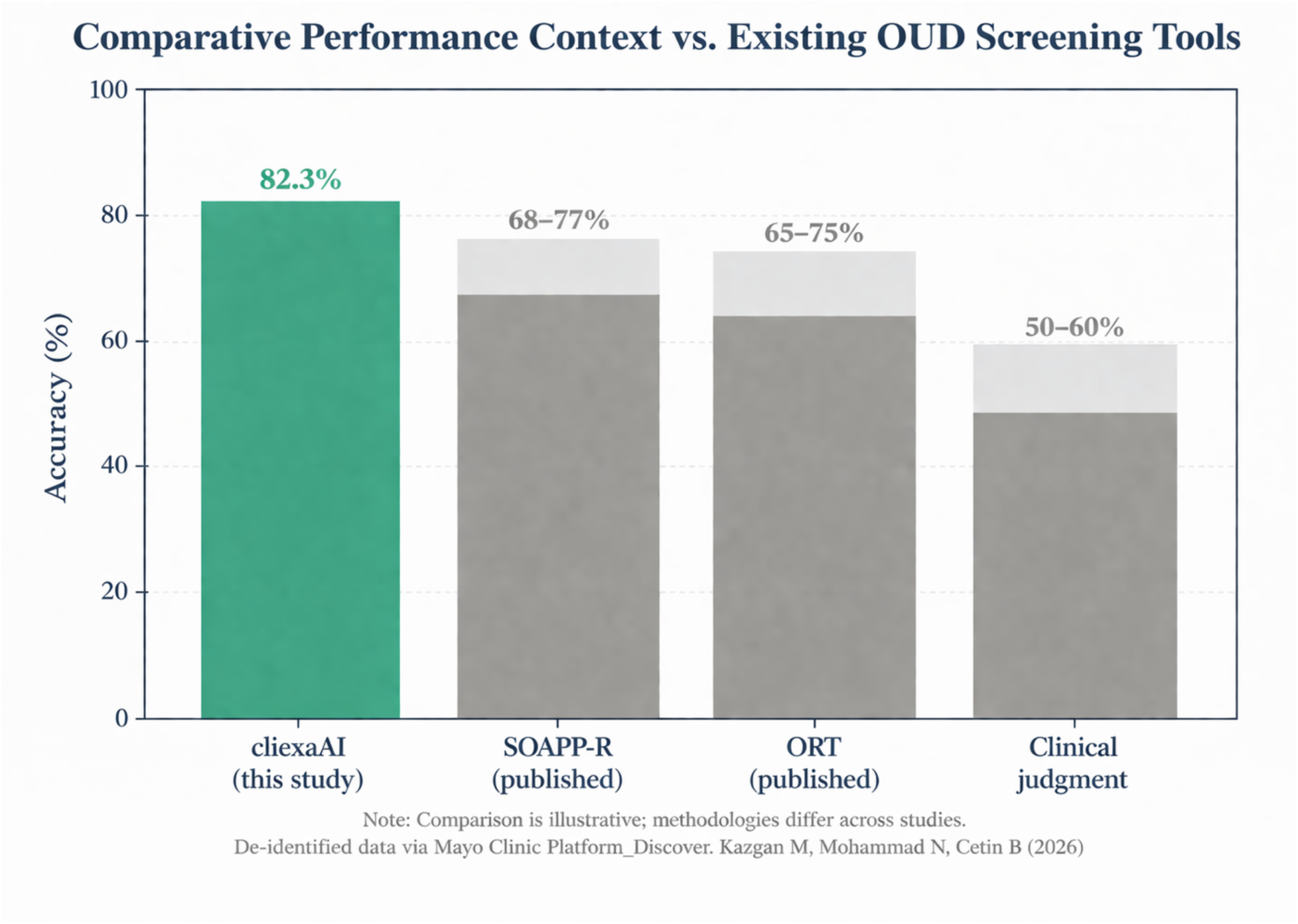
Comparative performance context versus existing OUD screening tools. The cliexaAI system’s 82.3% test set accuracy shown alongside published accuracy ranges for SOAPP-R (68–77%), ORT (65–75%), and clinical judgment alone (50–60%). Note: comparison is illustrative; methodologies differ across studies.

Beyond accuracy, the cliexaAI system offers structural advantages: it evaluates approximately 42 features across 6 clinical domains versus 5–24 items in self-report tools; provides four-tier gradation versus binary risk classification; supports continuous automated monitoring versus episodic screening; and delivers transparent risk factor attribution through the CRE rather than opaque composite scores.

## Discussion

This study presents the development and qualification of an explainable AI clinical decision support system for OUD risk prediction that addresses three critical gaps in the current landscape: the absence of dual-layer architectures combining ML with clinical reasoning, the lack of documented and measured bias mitigation in OUD prediction models, and the limited progression of such models from retrospective development to clinical platform qualification.

### The fairness–accuracy tradeoff

The central finding of our bias mitigation analysis is that a 1.9 percentage-point reduction in overall test accuracy (84.2% → 82.3%) purchased a 76% reduction in the racial performance gap for high-risk detection (30.3pp → 7.4pp) and a 59% reduction in age-based disparity (6.6pp → 2.7pp). We argue this tradeoff is not merely acceptable but clinically desirable. In a system designed to identify patients at risk for a potentially fatal condition, equitable detection across demographic groups is a safety requirement [10,19], not an optimization target.

The specific mechanisms driving this improvement age-balanced resampling, race feature removal, and cost-sensitive loss reweighting—are individually well-established in the fairness-ML literature [17,18]. Our contribution is demonstrating their combined application in a clinical context with measured outcomes, providing a quantified reference point for future work in responsible clinical AI development.

### Dual-layer architecture as a model for clinical AI

The combination of machine learning prediction (XGBoost) with rule-based clinical reasoning (CRE) addresses a fundamental tension in clinical AI deployment: ML models optimize for predictive accuracy, while clinical practice requires transparent reasoning that aligns with established diagnostic frameworks. The CRE serves three functions: (1) as an explainability layer that maps model outputs to interpretable clinical risk domains, (2) as a safety net that catches cases where ML prediction may diverge from clinical heuristics, and (3) as a calibration mechanism with clinician-derived tier boundaries that ensure risk categories carry consistent clinical meaning.

This architecture is intentionally not novel in its individual components. Its contribution lies in the integration pattern: a well-validated ML algorithm for pattern detection paired with a transparent, auditable rules engine for clinical interpretation. We propose this as a reproducible design pattern for clinical AI systems where both predictive power and explainability are required.

### The decision to exclude the MLP

Our initial development (V1) included a multi-layer perceptron (MLP) that achieved higher raw accuracy (90.5% vs. 84.2% on the original test set). We deliberately excluded it from the final submission for three reasons. First, the MLP achieved only modest racial gap improvement (30.3pp → 23.4pp) compared to the bias-mitigated XGBoost (30.3pp → 7.4pp), indicating that architectural complexity alone does not resolve demographic disparities. Second, XGBoost provides native feature importance metrics that integrate naturally with the CRE’s domain-based scoring, supporting the dual-layer architecture’s explainability goals. Third, tree-based models are more readily auditable and interpretable for clinical stakeholders, aligning with the responsible AI principles required for platform qualification.

This decision reflects a broader principle: in clinical AI, the best model is not always the most accurate model. It is the model that best balances accuracy, fairness, explainability, and deployability within the clinical context.

### Significance of platform qualification

The qualification of cliexaAI on Mayo Clinic Platform represents a meaningful validation step beyond traditional retrospective performance reporting. Mayo Clinic Platform_Solutions Studio evaluates not only algorithmic performance but also intended use definitions, clinical workflow integration, fairness evaluation methodology, documentation quality, and adherence to responsible AI principles. This multi-dimensional assessment provides greater assurance of clinical readiness than performance metrics alone.

### Clinical workflow implications

The system is designed to integrate into existing clinical workflows by providing real-time risk stratification within the EHR. By automating multi-factor risk assessment, it reduces reliance on manual screening tools and subjective clinical judgment. The four-tier risk output aligns with existing care pathways, enabling targeted interventions such as monitoring, referral, or treatment escalation.

## Limitations

Several limitations should be considered when interpreting these results.

First, the model was developed using de-identified data from a single institutional source accessed via Mayo Clinic Platform_Discover, with geographic concentration (77.3% from the west NorthEast region based on timez one analysis). Generalizability to other health systems, geographic regions, and patient populations requires external validation.

Second, the study uses a retrospective development design. While the model was qualified on Mayo Clinic Platform, prospective deployment with real-time clinical outcome tracking has not yet been conducted. The comparative performance context versus existing screening tools (SOAPP-R, ORT) is illustrative rather than definitive, as direct head-to-head evaluation was not performed. Taken together, this is a single-platform, internal-validation study without prospective clinical deployment; the present results should therefore be read as evidence of internal validity and of improved subgroup performance parity, not of real-world clinical effectiveness. We note candidly that this maturity stage is shared by much of the published clinical AI and CDSS literature, in which the large majority of models report only internal or retrospective validation and few progress to external or prospective evaluation [21]. Consistent with the FUTURE-AI Universality principle [22] and the TRIPOD+AI reporting guideline [23], external multi-site validation and prospective evaluation are widely recognized as prerequisites for clinical adoption, and we position them as the necessary next step for this work rather than as completed milestones.

Third, the High-risk tier represents only 7.0% of the balanced dataset (n=110 in the test set), limiting statistical power for this clinically critical category. Similarly, minority racial subgroups have limited representation (Black patients: 10.4% of balanced test set; Asian: 3.2%; Native American: 1.7%), constraining the precision of subgroup performance estimates.

Fourth, missing PRO data (∼12–15%) may affect risk stratification accuracy in real-world deployment where PRO completion rates may vary. The MAT-only patient exclusion may omit valid cases of opioid dependence, potentially introducing cohort misclassification.

Fifth, while SHAP-based per-patient explainability code has been developed, execution and integration into the clinical interface are planned for the next iteration. Current feature importance is based on aggregate gain metrics rather than individualized attribution.

Finally, the study does not evaluate downstream clinical outcomes such as prescribing behavior changes or reduction in OUD incidence, which will be addressed in future prospective studies.

## Conclusion

We present the development and qualification of an explainable AI clinical decision support system for opioid use disorder risk prediction. The dual-layer architecture combining XGBoost machine learning with a Clinical Rules Engine achieves 82.3% test accuracy with four-tier risk stratification while providing transparent, clinician-interpretable risk factor attribution. An iterative bias mitigation strategy reduced racial performance disparities by 76% and age-based disparities by 59%, demonstrating that equitable clinical AI is achievable through deliberate design choices at acceptable accuracy cost. The qualification on Mayo Clinic Platform establishes a pathway for responsible deployment of AI-driven decision support in addiction medicine. Future work will focus on prospective clinical evaluation, SHAP-based per-patient explainability, multi-site external validation, and temporal risk progression modeling.

## Data availability statement

This study involves analysis of de-identified data via Mayo Clinic Platform_Discover. In accordance with the Code of Federal Regulations, 45 CFR 46.102, the noted activity does not require IRB review. Data shown and reported in this manuscript has been extracted from this environment using an established protocol for data extraction, aimed at preserving patient privacy. The data has been de-identified pursuant to an expert determination in accordance with the HIPAA Privacy Rule. The underlying patient-level data cannot be shared publicly because they were accessed under the Mayo Clinic Platform_Discover data-use agreement and an expert-determination de-identification that prohibit redistribution; the authors do not have permission to release the raw data. Qualified researchers may request access to the de-identified data underlying the study by contacting cliexa, Inc. and Mayo Clinic Platform; access is subject to the governing data-use agreement and expert-determination terms. The model configuration and analysis code that do not reveal protected data are available from the corresponding author on reasonable request.

## Acknowledgments

The authors acknowledge the use of Mayo Clinic Platform_Discover for de-identified data access and Mayo Clinic Platform_Solutions Studio for independent solution qualification.

## Author contributions

Mehmet Kazgan conceived the study, designed the model architecture and supervised all aspects of development. Navid Mohammad contributed to data engineering, feature development, model selection, implementation, and bias mitigation strategy. Burak Cetin contributed to system architecture and Clinical Rules Engine development. All authors reviewed and approved the final manuscript.

## Supporting information

**S1 Table.** XGBoost hyperparameter configuration across model versions (V1–V3).

**S2 Table.** Test-set confusion matrix for the bias-mitigated model (V3).

